# Cardiac Output During Exercise: Thermodilution versus Direct Fick

**DOI:** 10.1101/2025.04.14.25325840

**Authors:** Matthew T. Siuba, James Lane, David Toth, Deborah Paul, Huijun Xiao, Xiaofeng Wang, Adriano R. Tonelli

## Abstract

**BACKGROUND:** Accurate cardiac output (CO) measurement during invasive cardiopulmonary exercise testing (iCEPT) is essential for classification of diseases such as exercise pulmonary hypertension (ex-PH), exercise pre-capillary PH (ex-prePH), and exercise postcapillary PH (ex-pcPH). The purpose of this study is to compare performance of thermodilution CO (tdCO) to gold-standard direct Fick CO (dfCO) during iCPET.

**METHODS:** A single-center prospective cohort study of patients undergoing iCPET over a three-year period. For the primary outcome, we predicted CO at each stage of exercise and recovery using generalized additive modeling. In secondary analysis, we assessed mean differences in CO across exercise stages. Finally, we assessed the impact in classification of ex-PH, ex-prePH, and ex-pcPH between the two CO methods.

**RESULTS:** A total of 302 patients were included. In the primary analysis, the generalized additive model smooth term was significantly different between tdCO and dfCO (p < 0.001), with tdCO underestimating dfCO at rest and overestimating dfCO at max exercise. Mean differences between tdCO and dfCO were only significantly different across two stages of exercise. Furthermore, classification rate of ex-PH, ex-prePH, and ex-pcPH was not significantly different between cardiac output methods (4, 3, and 1 patient misclassified by tdCO, respectively).

**CONCLUSION:** While there are differences in CO estimation between thermodilution and direct Fick during iCPET, the degree of error did not meaningfully affect the hemodynamic classification of patients with ex-PH, ex-prePH, or ex-pcPH.

## Introduction

Exercise right heart catheterization (RHC) is commonly performed to evaluate patients with unexplained dyspnea and/or exercise intolerance. This test is essential to diagnose exercise pulmonary hypertension (ex-PH), exercise postcapillary PH (ex-pcPH), and preload insufficiency.^1^ For all these diagnoses, an accurate cardiac output (CO) measurement is essential both at rest and during exercise, since it allows for the determination of mean pulmonary artery pressure (mPAP)/CO and pulmonary artery wedge pressure (PAWP)/CO slopes as well as the peak CO percentage of predicted. In addition, CO measurement allows for the calculation of cardiac index, stroke volume, and pulmonary vascular resistance (PVR) at rest and during exercise.

The gold standard for CO assessment is the direct Fick CO (dfCO) method, which requires the measurement of oxygen consumption with a metabolic cart and arterial and venous oxygen content, under the same conditions. For the arterial and venous oxygen content, both arterial and mixed venous blood gas need to be simultaneously obtained and analyzed with co-oximetry with their respective measurement of hemoglobin. Given this complexity, CO is commonly determined using thermodilution (tdCO), a methodology that is widely available and can be easily used during rest and exercise.

At rest, tdCO has high accuracy compared to dfCO, but wide limits of agreement.^2^ However, at rest, this tdCO determination is obtained in triplicate with measurements recorded only if there is < 10% variation. This methodologic requirement for tdCO is not possible during exercise since the time limitations of each exercise stage / condition reduce the ability to repeat tdCO both in ramp and step protocols. We previously contrasted CO methodologies during exercise in a smaller number of patients, and noted that tdCO compared to dfCO had wide limits of agreement during exercise, but similar predicted CO using a generalized additive model.^3^

Whether tdCO is sufficiently accurate to be used during exercise RHC remains unknown. Therefore, we tested this question in a large prospective cohort of patients who underwent upright invasive cardiopulmonary exercise testing (iCPET) with simultaneous measurement of tdCO and dfCO at rest and at every stage of exercise and recovery. Given the inherent limitations of tdCO and the fact that determinations are usually single given time constrains of exercise stages, we hypothesized that tdCO would be inaccurate during exercise with wide limits of agreement and this may have implications for the diagnosis of ex-PH, ex-pcPH, and ex-precapillary PH, conditions described by slopes or calculations that include CO.

## Methods

### Study design, participants, and setting

We performed a single-center, prospective study, in consecutive patients referred for iCPET from March 2021 to August 2023. The iCPET adds to a traditional exercise RHC a radial arterial line and CPET ^4^. The radial line allows for the simultaneous acquisition of ABG and VBG from the pulmonary artery. The CPET provides important measurements including the oxygen consumption at the time of ABG and VBG acquisition. A subset of these patients (n = 24) were included in a previous study that tested the value of a non-invasive methodology, i.e. pulse contour analysis, in CO determination during exercise.^3^ The current study was approved by the institutional review board (#16-872) and informed consent was obtained from every patient.

Findings are reported following the Strengthening the Reporting of Observational Studies in Epidemiology (STROBE) guidelines.^5^

### Exercise protocol

Patients underwent maximal exercise in a sitting cycle ergometer, pedaling at a cadence of 60 revolutions/minute using a step protocol with increments of 20 Watts (W) every 2 minutes. Hemodynamic determinations including right atrial pressure, pulmonary artery pressure, and pulmonary artery wedge pressure were obtained at all stages. Pulmonary pressures were averaged across the respiratory cycle to account for pressures swings associated with changes in intrathoracic pressure during exercise.

At baseline, in upright position, tdCO was performed in triplicate to obtain a CO within 10% of one another. Due to limited duration of the exercise stages (2 minutes) and all the determinations involved (pulmonary pressure and VBG), we obtained only one tdCO measurement during each stage of exercise. The VBG is obtained from the distal port of the pulmonary artery catheter, and after this blood is collected, flushing with normal saline is needed to avoid clotting and pressure waveform damping. This flushing makes the temperature sensor unstable and not ready for thermodilution. For unusual cases (< 5%) in which CO was inconsistent with the expected increase during exercise (e.g. increase by more than 50% or a decrease), the measurement was repeated, and the most reasonable CO value kept. Recovery was assessed at one- and three-minute post exercise.

At baseline and at every stage of exercise and recovery, we recorded tdCO. Immediately after recording tdCO, 2 nurses simultaneously obtained ABG and VBG after discarding 3 mL of blood, to account for 3 times the dead space between the tip of the catheter and the collection syringe (Portex, Smiths Medical ASD Inc, MN, USA). Extra care was taken to remove any bubble of air, to prevent falsely elevated oximetric determinations, and to turn/rotate the syringe to prevent clotting. ABG and VBG were immediately sent to the laboratory using a pneumatic tube transport system. Blood was immediately processed using a blood gas analyzer with co-oximetry (Radiometer America ABL800 Flex+ system, CA, USA). At the same time of obtaining ABG and VBG, we carefully recorded the oxygen consumption (Ultima CPX, MGC Diagnostics, Saint Paul, MN). A detailed explanation of our iCPET protocol was described in a prior publication.^6^

### Statistical analysis

Continuous variables are presented as median (interquartile range [IQR]) and categorical variables as counts (percentages) unless otherwise specified. Paired t-tests were used to assess mean differences in tdCO and dfCO. We fitted generalized additive models (GAMs) for both CO methods, to assess the primary aim of this study - the difference between tdCO and dfCO across all stages of exercise, including baseline and recovery -. We fit a GAM for both CO methods across exercise stages, since we expected non-linear trends in measurements as previously observed.^3,7^ A random effect was included to adjust for patient-specific variability. Exercise stages were mapped onto a numerical scale ranging from 0 (baseline) to 1 (maximum stage), with intermediate stages evenly spaced. This scaled was used to normalize the variable number of exercise stages across patients. Recovery stages were standardized to 1.25 for 1-minute recovery and 1.5 for 3-minute recovery, assuming equal-spacing between intervals. The GAM model employed does not treat the baseline CO values as the mean of the baseline data points. Instead, it uses a smoothing function to capture the overall relationship between each CO and exercise stage for every patient. Therefore, the GAM prediction is not simply a summary of baseline data but an estimate of how baseline CO fits into the overall relationship between CO and exercise stages.

Secondary analyses included mean differences between CO methods at baseline and across the first four stages of exercise (since this intensity of exercise was achieved by most patients), as well as the maximum exercise stage. We also report on the impact of tdCO determination in final diagnosis of ex-PH, ex-pcPH and exercise pre-capillary PH, considering dfCO as the gold standard. We computed slopes for mPAP/CO and PAWP/CO using all exercise stages. A mPAP/CO slope > 3 WU was used to diagnose ex-PH and a slope of PAWP/CO > 2 WU was used to diagnose ex-pcPH. A PVR > 2 WU at peak exercise was used to identify exercise precapillary PH (ex-prePH). A two-sided alpha of < 0.05 was considered significant. All analyses were performed using R Statistical Software (R Foundation for Statistical Computing, Vienna, Austria).

## Results

### Patient characteristics

A total of 302 patients were enrolled, 202 (67%) female, median age 49 (IQR 37, 63) years, with BMI 30.24 (IQR 25.96, 36.59) kg/m^2^ (**Table 1)**. No patients had more than mild tricuspid regurgitation on echocardiography. A summary of hemodynamic data at baseline and across all exercise stages is included in **Table 2**. There was no missing data across exercise stages.

**Table 1.** Key Patient Characteristics.

| Characteristic | N = 302 |
| --- | --- |
| Age (years) | 49 (37,63) |
| Female (%) | 202 (67%) |
| Height (cm) | 167.6 (162.6, 175.3) |
| Weight (kg) | 87.5 (72.9, 103) |
| Body surface area (BSA) (m<sup>2</sup>) | 1.99 (1.80, 2.13) |
| Body mass index (BMI) (kg/m<sup>2</sup>) | 30.24 (25.96, 36.59) |
| Resting PH | 37 (12.3%) |
| Post-procedure diagnoses (n = 265)* |  |
| Preload insufficiency | 132 (49.8%) |
| Exercise precapillary PH | 43 (16.2%) |
| Exercise PH | 41 (15.5%) |
| Exercise postcapillary PH | 4 (1.5%) |
| Median (IQR) unless otherwise specified. *Patients without resting pulmonary hypertension |  |

**Table 2.**
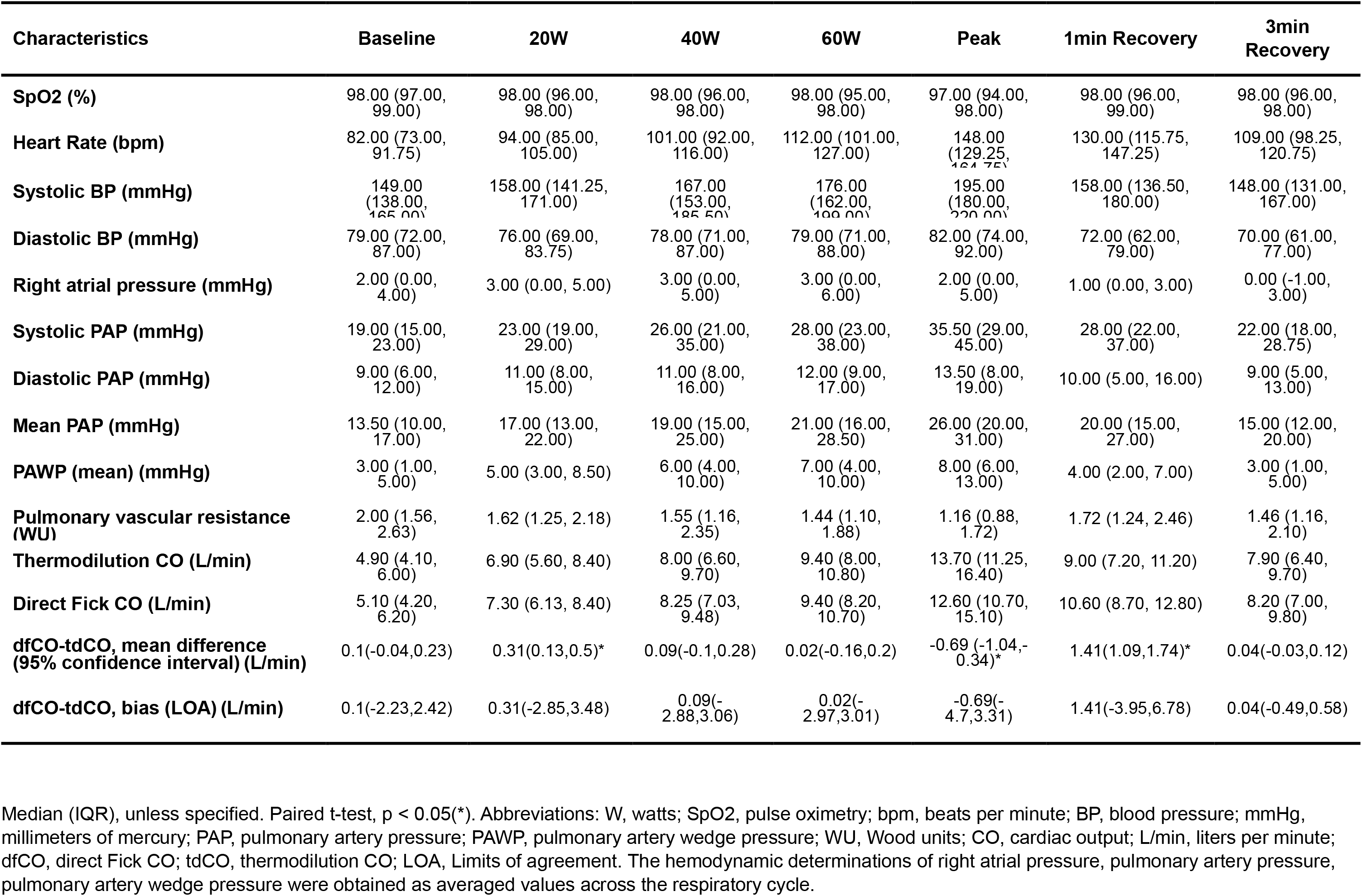
Hemodynamics by stage.

Hemodynamic phenotype on iCPET

Resting PH (mPAP > 20 mmHg) was present in 37 (12.3%) patients. Of the remaining 265 patients, 132 (49.8%) were diagnosed with preload insufficiency, 43 (16.2%) with ex-prePH, 41 (15.5%) ex-PH, and 4 (1.5%) with ex-pcPH (**Table 1)**.

### Comparison between tdCO and dfCO

We noted significant differences in smooth terms between tdCO and dfCO with GAMs (p < 0.001) (**Figure 1)**. Specifically, tdCO overestimates dfCO at rest and underestimates dfCO at peak exercise. The predicted increase in dfCO per 20W was 0.45 L/min (SD 1.50 L/min). The expected decrease in dfCO from peak to 1 and 3 min recovery was 1.25 L/min and 3.41 L/min, respectively.

**Figure 1.**
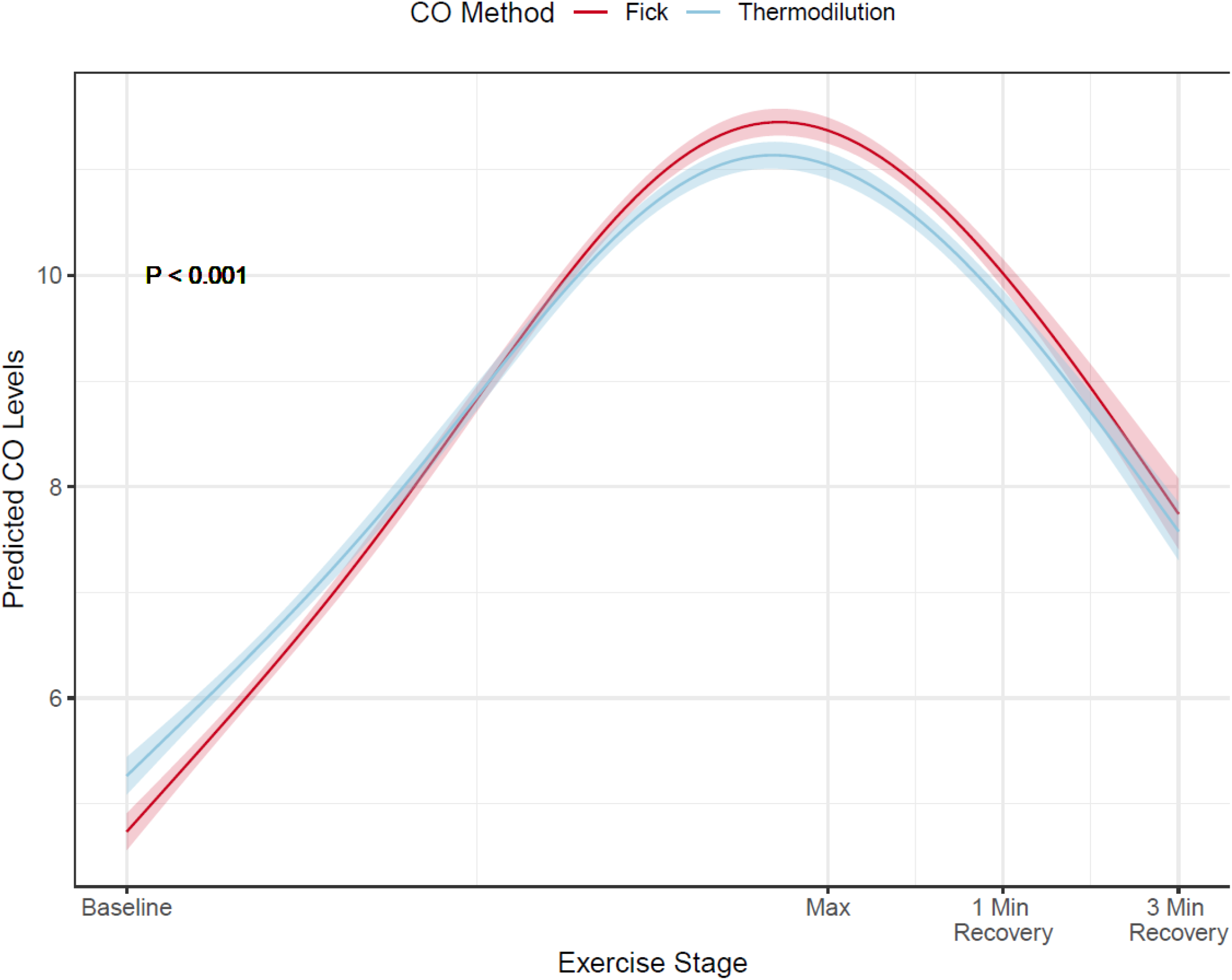
Generalized additive model for comparison of cardiac output methods.

Mean differences and limits of agreement (LOA) for dfCO and tdCO across exercise stages and recovery are shown in **Table 2**. Mean differences were only statistically significant across two stages of exercise (20W and peak exercise) as well as 1 min recovery. The LOA in CO between dfCO and tdCO ranged from the lowest at 3 min of recovery (−0.49, 0.58 L/min), to largest at 1 min of recovery (−3.95, 6.78 L/min).

### Secondary outcomes

In the subset of 265 patients with normal resting pulmonary pressures, the mean mPAP/dfCO slope was 1.99 WU compared to mean mPAP/tdCO slope of 2.18 WU (mean difference −0.18, 95% CI −0.25, −0.13, p < 0.001). The mean PAWP/dfCO slope was 0.68 WU compared to mean PAWP/tdCO slope of 0.71 WU (mean difference −0.03, 95% CI −0.05, 0.002, p = 0.07). PVR at max exercise was 1.65 (SD 1.26) WU using dfCO and 1.70 (SD 1.36) WU with tdCO, a non-significant difference (mean difference −0.01, 95% CI −0.06, 0.03, p = 0.6).

Using dfCO, 43 patients were classified as ex-prePH compared to 46 patients with tdCO, 41 (15.5%) of patients were classified as ex-PH compared to 45 (17.0%) with tdCO. Four patients (1.5%) were classified as ex-pcPH using dfCO compared to five (1.9%) with tdCO **(Table 3)**.

**Table 3.**
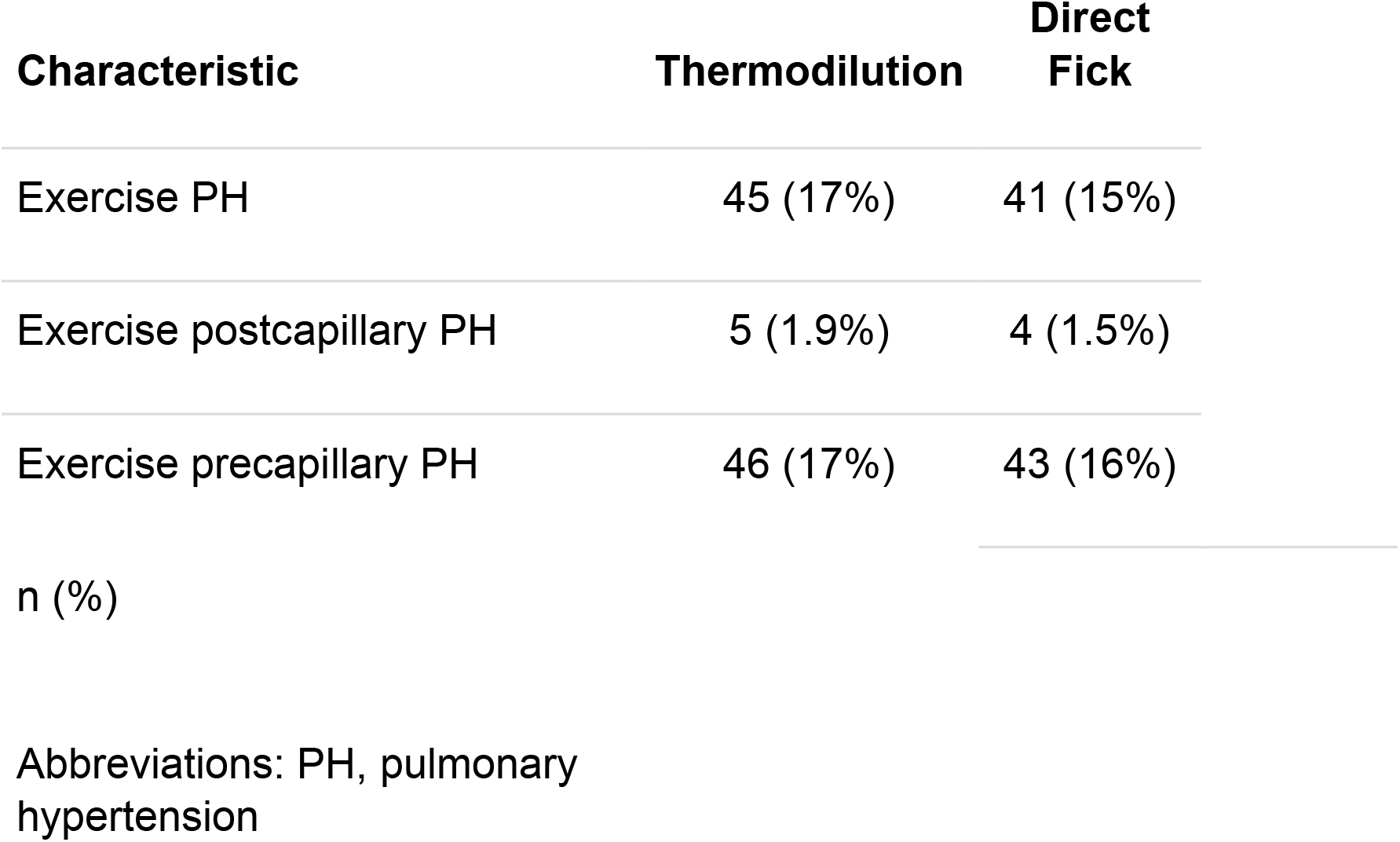
Hemodynamic Exercise Classification by Cardiac Output Methodology.

| Characteristic | Thermodilution | Direct Fick |
| --- | --- | --- |
| Exercise PH | 45 (17%) | 41 (15%) |
| Exercise postcapillary PH | 5 (1.9%) | 4 (1.5%) |
| Exercise precapillary PH | 46 (17%) | 43 (16%) |
| n (%) |  |  |

Abbreviations: PH, pulmonary hypertension

Comparison of classification by CO method is shown in **Figure 2**.

**Figure 2.**
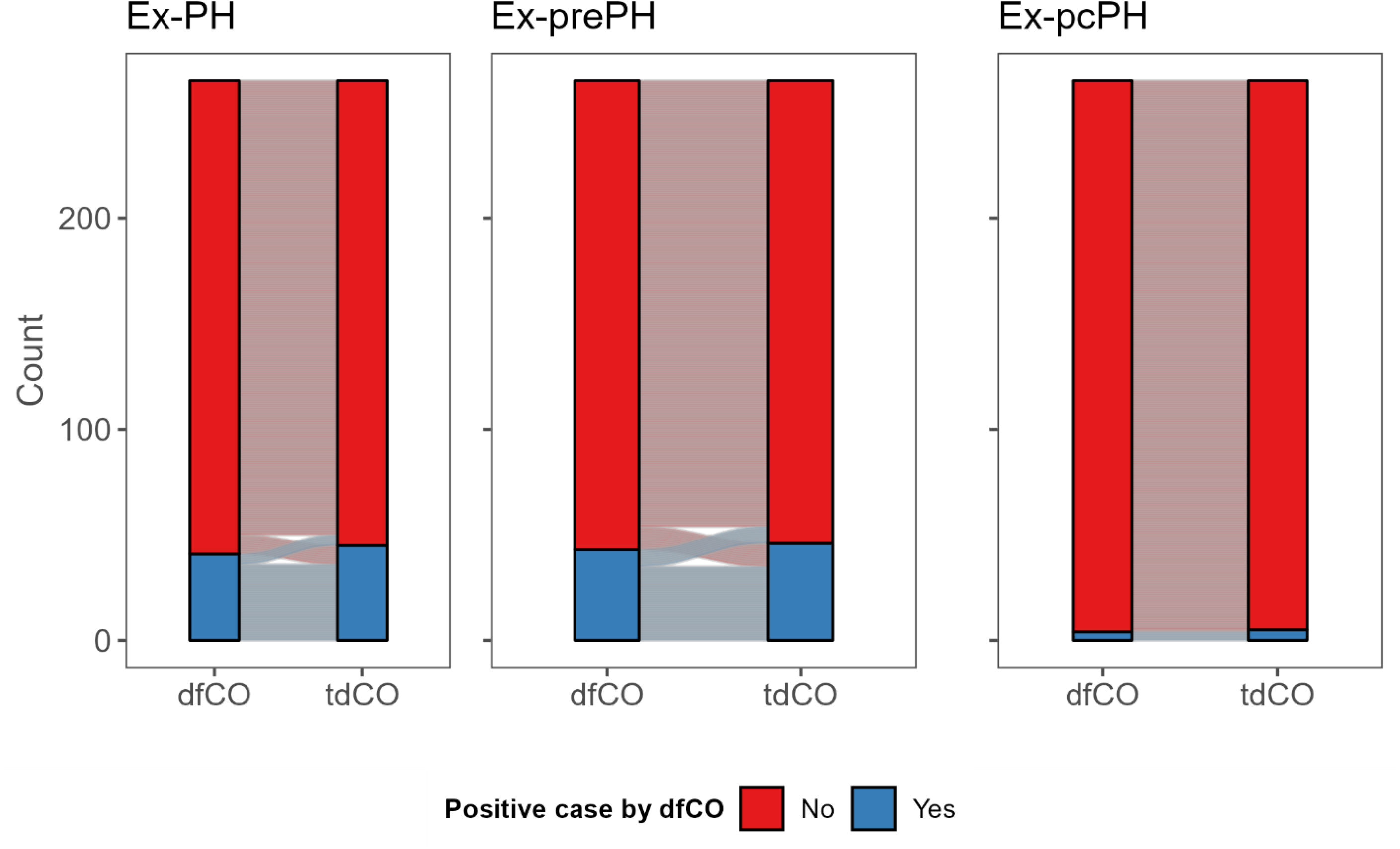
Classification of Exercise Phenotypes by Cardiac Output Method. Abbreviations: Ex-PH, exercise pulmonary hypertension; ex-prePH, exercise precapillary pulmonary hypertension; Ex-pcPH, exercise postcapillary pulmonary hypertension; dfCO, direct Fick cardiac output; tdCO, thermodilution CO.

## Discussion

In the largest study of its kind to date, we demonstrated significant differences between dfCO and tdCO during exercise using an advanced modeling approach. We noted a higher tdCO at rest and lower at peak exercise when compared to the gold standard dfCO. Additionally, there was a statistically significant difference between mPAP/CO slopes between tdCO and dfCO, and a borderline difference between PAWP/CO slopes, that did not meaningfully alter the classification of patients with ex-PH and ex-pcPH. The PVR at peak exercise was numerical lower when using dfCO (although not significantly different), and this small variation misclassified a few patients with PVR > 2 WU at peak exercise.

This study differs from prior evaluations of tdCO and dfCO during exercise in the large number of patients included, the contemporary, rigorous and sound methodology used, the absence of missing values, and the inclusion of a cohort in whom the majority had normal resting hemodynamics. Hsu et al^8^ reported a larger difference in mPAP/CO slopes (roughly 1 mmHg/l/min higher in the tdCO group) and maximal exercise cardiac outputs (2.3 L/min lower in tdCO), which lead to a higher rate of misclassification of patients with ex-PH. However, their mean exercise slopes were closer to the mPAP/CO discrimination value of 3 WU for ex-PH than our cohort. In addition, this study included 20 patients with normal resting hemodynamics, compared to 265 patients in our study, and used supine bicycle ergometry rather than upright bicycle as in our work. Our group previously demonstrated differences in supine and upright hemodynamics during exercise, such as filling pressures and CO, though differences in exercise mPAP/CO and PAWP/CO slopes were not detectably different.^9^ Similar to the Hsu study, we identified wide limits of agreement between tdCO and dfCO at several stages.^3,8^

Although there was a numerical difference in tdCO and dfCO, this difference had only minor clinical implications, a finding that is very important to support the regular use of tdCO determinations during exercise which is a much more accessible methodology. These findings, particularly when considering the most patients only had one determination of dtCO per stage, is likely due to the use of mPAP/CO and PAWP/CO slopes, that allow for correction of intrinsic variations in the measurement of tdCO during exercise. A specific value may be higher or lower than the gold standard dfCO; however, when these measurements are repeated there is regression to the mean, minimizing the measurement error. However, then tdCO is taken a specific point and used in calculations such as baseline and peak exercise PVR, the differences are more pronounced with direct clinical implications.

Interestingly, the impact of the CO methodology on the exercise hemodynamic classification depends on the LOA of tdCO with the standard dfCO and the characteristics of the cohort investigated. Variations in tdCO compared to dfCO have more pronounced implications when PVR at peak exercise and mPAP/CO and PAWP/CO are close to the discrimination points used to define ex-PH, ex-pcPH and ex-prePH. Therefore, clinicians should be particularly careful when key calculations using tdCO are closer to the cut-off, particularly when only one specific measurement of tdCO is used, like in the calculation of PVR. Peak PVR may be more susceptible to errors in tdCO, since this measurement cannot be usually repeated.

It is important to assess tdCO measurements both at rest and exercise and repeat (if possible) or critically examine and value (if repetition is not possible) measurements that are discordant with prior or expected determinations. For example, if tdCO decreases with exercise or if the increase is markedly disproportional from one stage to the other, then this measurement should be repeated, even if the stage of exercise is slightly prolonged. To reduced errors tdCO requires careful determination, including a specific volume of injection (usual 10 milliliters) at a constant temperature with steady injection and adequate programming of the measuring system, selecting the appropriate volume of injection and computation constant (or brand and type of pulmonary artery catheter used if already part of the software).

The determination of a reliable dFCO also requires important steps and precautions.

These include simultaneous acquisition of ABG and VBG by two operators, as oxygenation may change during exercise or other conditions. Pulse oximetry cannot be used, given large LOA with arterial oxygen saturation.^10^ Both ABG and VBG need to be done with co-oximetry to eliminate the impact of carboxy- and methemoglobin in the percentage of oxyhemoglobin.^10,11^ ABG and VBG should have no air in the syringe as this will affect oximetric determinations^12^ and need to be adequately rotated for an adequate heparin distribution to prevent clotting that would render a blood gas pair unusable. We use the hemoglobin reported on the VBG in each stage, since this value may vary during exercise. Oxygen consumption should be measured at the time of blood gases collection, following established criteria and averaging values over several seconds to reduce errors in the measurement.

This study has limitations, including its single-center nature. While there was no significant misclassification in ex-PH and ex-pcPH between CO methods, this study may have been underpowered to detect small differences, given the relatively low prevalence of these conditions in our cohort. The very low number of patients with ex-pcPH likely reflects the referral base for our iCPET which predominantly serves patients with unexplained dyspnea and exercise intolerance, which in great proportion have normal resting pulmonary hemodynamics. We did not specifically record the times in which tdCO was repeated during exercise due to unreliable determinations (e.g. drop in CO or unexpected increases), but authors estimated that it was around 5% of the measurements. Future study in this area could be enriched by including patients with higher pre-test probability for one of these exercise-induced conditions. Despite these limitations, this study provides support to use thermodilution for CO determination during exercise, particularly when using mPAP/CO and PAWP/CO slopes and repeating unexpected values.

## Conclusion

While there are differences in CO estimation between thermodilution and direct Fick during exercise, this did not meaningfully affect the hemodynamic classification of patients with ex-PH, ex-prePH, or ex-pcPH and supports the use of thermodilution for regular exercise RHC.

## Abbreviation List

BMI: body mass index
BP: blood pressure
CI: confidence interval
CO: cardiac output
CPET: cardiopulmonary exercise test
DF: direct Fick
Ex-pcPH: exercise postcapillary pulmonary hypertension
Ex-PH: exercise pulmonary hypertension
Ex-prePH: exercise precapillary pulmonary hypertension
GAM: generalized additive model
IQR: interquartile range
mmHg: millimeters of mercury
mPAP: mean pulmonary artery pressure
PA: pulmonary artery
PAWP: pulmonary artery wedge pressure
PH: pulmonary hypertension
PVR: pulmonary vascular resistance
RHC: right heart catheterization
TD: thermodilution
W: Watts
WU: Woods unit

## Data availability statement

The data underlying this article cannot be shared publicly due to the limitations imposed by the institutional review board. The data will be shared on reasonable request to the corresponding author after creating a data use agreement between institutions.

## Contributions

MTS participated in data analysis, drafted the original manuscript, participated in revision. MGS, JL, DP participated in data curation, patient enrollment, and critical revision of the manuscript. HX and XW participated in data analysis and critical revision of the manuscript. ART conceived the study and participated in data interpretation, drafting and critical revision of the manuscript and is the guarantor of the manuscript. All gave final approval and agreed to be accountable for all aspects of work ensuring integrity and accuracy.

## References

1. Montané B, Tonelli AR, Arunachalam A, et al. Hemodynamic Responses to Provocative Maneuvers during Right Heart Catheterization. Ann Am Thorac Soc. 2022;19(12):1977–1985. doi:10.1513/AnnalsATS.202201-077OC

2. Khirfan G, Ahmed MK, Almaaitah S, et al. Comparison of Different Methods to Estimate Cardiac Index in Pulmonary Arterial Hypertension. Circulation. 2019;140(8):705–707. doi:10.1161/CIRCULATIONAHA.119.041614

3. Siuba MT, Sharpe MG, Lane J, et al. Cardiac output measurement during invasive cardiopulmonary exercise testing: comparing pulse contour analysis, thermodilution, and Fick methodology. Eur J Prev Cardiol. 2024;31(12):1550–1552. doi:10.1093/eurjpc/zwae143

4. Tooba R, Mayuga KA, Wilson R, Tonelli AR. Dyspnea in Chronic Low Ventricular Preload States. Ann Am Thorac Soc. 2021;18(4):573–581. doi:10.1513/AnnalsATS.202005-581CME

5. von Elm E, Altman DG, Egger M, et al. The Strengthening the Reporting of Observational Studies in Epidemiology (STROBE) statement: guidelines for reporting observational studies. J Clin Epidemiol. 2008;61(4):344–349. doi:10.1016/j.jclinepi.2007.11.008

6. Nanah A, Garcia MVF, Lane J, Paul D, Tonelli AR. Plasma catecholamines in patients undergoing invasive cardiopulmonary exercise test for exercise intolerance. Respir Med. 2024;233:107775. doi:10.1016/j.rmed.2024.107775

7. Hastie T, Tibshirani R. Generalized additive models for medical research. Stat Methods Med Res. 1995;4(3):187–196. doi:10.1177/096228029500400302

8. Hsu S, Brusca SB, Rhodes PS, Kolb TM, Mathai SC, Tedford RJ. Use of thermodilution cardiac output overestimates diagnoses of exercise-induced pulmonary hypertension. Pulm Circ. 2017;7(1):253–255. doi:10.1086/690629

9. Kirupaharan P, Lane J, Melillo C, et al. Impact of body position on hemodynamic measurements during exercise: A tale of two bikes. Pulm Circ. 2024;14(1):e12334. doi:10.1002/pul2.12334

10. Ascha M, Bhattacharyya A, Ramos JA, Tonelli AR. Pulse Oximetry and Arterial Oxygen Saturation during Cardiopulmonary Exercise Testing. Med Sci Sports Exerc. 2018;50(10):1992–1997. doi:10.1249/MSS.0000000000001658

11. Khirfan G, Ahmed MK, Faulx MD, Dakkak W, Dweik RA, Tonelli AR. Gasometric gradients between blood obtained from the pulmonary artery wedge and pulmonary artery positions in pulmonary arterial hypertension. Respir Res. 2019;20(1):6. doi:10.1186/s12931-018-0969-7

12. Biswas CK, Ramos JM, Agroyannis B, Kerr DN. Blood gas analysis: effect of air bubbles in syringe and delay in estimation. Br Med J Clin Res Ed. 1982;284(6320):923–927. doi:10.1136/bmj.284.6320.923

